# Healthcare Cost of Stargardt Disease

**DOI:** 10.1101/2020.01.22.20017962

**Authors:** Kanza Aziz, Bonnielin K. Swenor, Joseph K. Canner, Mandeep S. Singh

## Abstract

**Importance:** Stargardt disease (SD) is the most common juvenile macular degeneration and a leading cause of uncorrectable childhood blindness. The progressive and incurable nature of this chronic condition entails a long-term financial burden on affected individuals. The economic costs of SD have not been characterized in detail.

**Objective:** To estimate the direct healthcare cost of SD.

**Design:** Cross-sectional analysis of healthcare claims.

**Participants:** Patients with an ICD-9 diagnosis code of SD, non-exudative age-related macular degeneration (AMD), or bilateral sensorineural hearing loss (SHL).

**Methods:** Outpatient administrative claims data from the IBM® MarketScan® Commercial Claims and Encounters Database from 2010 to 2014 were analyzed.

**Main Outcome Measure:** Annual per-patient direct healthcare cost.

**Results:** A total of 472,428 patients were analyzed (5,015 SD, 369,750 SHL and 97,663 AMD patients respectively). The payment per year of insurance coverage for SD (median: $105.58, IQR: $50.53-$218.71) was higher than that of SHL (median: $51.01, IQR: $25.66-$121.66, p <0.001) and AMD (median: $76.20, IQR: $38.00-$164.86, p <0.001). When adjusted for covariates, the annual payment for SD was $47.83 higher than SHL (p<0.001) and $17.34 higher than AMD (p<0.001).

**Conclusions and Relevance:** There is a significant direct healthcare cost associated with SD. The annual per-patient cost of SD was higher than SHL, another condition that causes sensory impairment in people of all ages, and nonexudative AMD which causes a similar pattern of visual loss that typically begins later in life. The total lifetime per-patient cost of SD may exceed that of nonexudative AMD.

## Introduction

Stargardt disease (SD) is the most common form of inherited macular degeneration, and is caused by mutations in the *ABCA4* gene. Its reported incidence is 1 in 8,000 - 10,000^1^. People with SD typically begin to experience progressive visual loss as children or young adults^2^. Although there is no cure at present, several possible treatments including pharmacologic, stem cell, and gene therapy modalities are being evaluated in clinical trials^3,4^. Without treatment, vision loss in SD is typically progressive. Approximately one decade after symptom onset, visual acuity typically reaches the 20/100 to 20/200 level^5,6^. Management strategies for symptomatic patients currently include disease monitoring, counseling, and low vision rehabilitation. When coupled with the current lack of treatment options, the progressive nature of SD entails long-term healthcare-related financial implications for patients. Furthermore, available data suggest that early-onset SD tends to run a more severe course, and causes severe permanent vision loss earlier in life, than later-onset SD^3,7^. Those affected by earlier SD onset may require more medical care and therefore incur greater costs.

There is a paucity of information on the healthcare costs of SD. Presumably this is because SD is regarded as a rare disease, unlike age related macular degeneration (AMD)^8,9^. The global AMD treatment market is expected to reach $11.6 billion by 2026^10^. SD is currently the target of therapeutic development; estimates of the healthcare costs associated with SD would be useful in determining the total impact of these therapies. Healthcare costs have significant implications for both the individual and society because these costs constitute an important driving factor behind healthcare policy and economics, especially for chronic diseases such as SD.

To better understand healthcare costs associated with SD in the context of other disease conditions, we also analyzed the costs associated with non-exudative AMD and bilateral sensorineural hearing loss (SHL) as controls. The rationale for selecting non-exudative AMD was that the typical pattern of vision loss during the course of disease is analogous to SD to some degree. The macula is the site of retinal cell degeneration in both these conditions. Therefore, central, rather than peripheral, vision loss is progressively compromised over time, and is the chief source of vision-related disability and healthcare-seeking behavior in both conditions. We selected SHL as another control because it is the impairment of a sensory system, other than vision, that affects younger individuals and entails long-term healthcare cost implications. Therefore, the aim of this study was to estimate the direct healthcare costs of SD as compared to non-exudative AMD and SHL.

## Methods

### Data Source

Outpatient insurance claims data from 2010 to 2014 were analyzed using the IBM® MarketScan® Commercial Claims and Encounters Database. MarketScan® is a large database containing adjudicated and paid insurance claims data of individuals and their dependents whose health insurance is provided by their employers. These include active employees, early retirees and Consolidated Omnibus Budget Reconciliation Act (COBRA) continuers. Most of the data are collected from large employers and health plans. The types of benefit plans include comprehensive, exclusive provider organization (EPO), health maintenance organization (HMO), preferred provider organization (PPO), consumer directed health plan (CDHP), high deductible health plan (HDHP) and point-of-service (POS) plans with and without capitation. A unique identifier assigned to each patient is coded on claims. The dataset complies with Health Insurance Portability and Accountability Act regulations for a limited data-set and has undergone a third party review for a fully de-identified dataset^11^. The study was approved by the Johns Hopkins Medicine Institutional Review Board.

### Study Population

Claims with a primary diagnosis of SD, SHL and AMD were selected using ICD-9 diagnosis codes 362.75, 389.18 and 362.51, respectively. Claims with missing procedure codes were excluded. The unique patient identifier coded on claims was used to associate claims to each patient. In cases where a patient had more than one diagnosis, the primary diagnosis was used for categorization. Patients aged <50 years with a diagnosis of AMD were assumed to have been misdiagnosed or mis-coded and so were excluded from analysis.

SHL patients with cochlear implants were excluded due to lack of a comparable device to improve function in SD. Patients with exudative AMD, cystoid macular degeneration, macular cyst, hole or pseudohole, macular pucker or toxic maculopathy were excluded.

### Statistical Analyses

The gross median payments to providers for health services per year of insurance coverage were calculated for SD, SHL and AMD. We accounted for periods of non-coverage by considering only the total number of insured days for each patient.

Categorical variables were reported as frequencies and percentages while continuous variables were reported as medians with interquartile ranges. The Wilcoxon rank-sum test was used to test for statistical differences in the number of insurance claims per covered year and payments per covered year between groups. A multivariable quantile regression was used to adjust for covariates and analyze payments per covered year for each condition. Furthermore, a multivariable quantile regression model was used to compare payments per covered year between SD and SHL. Additional multivariable regression models were used to compare payments per covered year for SD with patients restricted to those aged ≥50 years and AMD for a more effective comparison. Covariates selected for the regression models included age (continuous variable), gender (male or female), year of first service in the database (2010-2014) and type of benefit plans (PPO, comprehensive, HMO, PPO with and without capitation, POS, CDHP and HDHP). The covariates were categorized as in the database.

A two-tailed P value of <0.05 was considered to be statistically significant. All statistical analyses were performed using Stata/MP version 14.2 (StataCorp LLC, College Station, TX).

## Results

### Population Characteristics

A total of 472,428 patients were analyzed (Table 1). Of these were 5,015 (1.1%) patients with SD, 369,750 (78.3%) with SHL, and 97,663 (20.7%) with AMD. Patients with SD (median age: 41 years, IQR [interquartile range]: 23-53 years) were younger than those with SHL (median age: 54 years, IQR: 44-60 years, p <0.001), and AMD (median age: 60 years, IQR: 56-62 years, p <0.001). The majority of SD and AMD patients were female (55.0% and 58.9%, respectively), whereas the majority of SHL patients were male (53.0%). The mean total number of gaps in insurance coverage per patient was 0.13 ± 0.41.

**Table 1.** General characteristics of the study population

| **Characteristics** | **SD**<br>(N=5,015) | **SHL**<br>(N=369,750) | **AMD**<br>(N=97,663) |
| --- | --- | --- | --- |
| **Age, median (IQR) yrs** | 41 (23 - 53) | 54 (44 - 60) | 60 (56 - 62) |
| **Sex, no. (%)** |  |  |  |
| **Male** | 2,257 (45.00) | 196,011 (53.01) | 40,186 (41.15) |
| **Female** | 2,758 (55.00) | 173,739 (46.99) | 57,477 (58.85) |
| **Type of benefit plan, no. (%)** | n= 4,600 | n=334,156 | n=87,576 |
| PPO | 98 (2.13) | 8,891 (2.66) | 3,574 (4.08) |
| Comprehensive | 157 (3.41) | 9,026 (2.70) | 2,228 (2.54) |
| EPO | 538 (11.70) | 42,979 (12.86) | 7,918 (9.04) |
| HMO | 336 (7.30) | 25,916 (7.76) | 7,211 (8.23) |
| POS | 3,036 (66.00) | 215,970 (64.63) | 59,372 (67.79) |
| POS with capitation | 28 (0.61) | 2,084 (0.62) | 605 (0.69) |
| CDHP | 209 (4.54) | 16,947 (5.07) | 3,965 (4.53) |
| HDHP | 198 (4.30) | 12,343 (3.69) | 2,703 (3.09) |

SD: stargardt disease, SHL: bilateral sensorineural hearing loss, AMD: non-exudative age related macular degeneration, IQR: interquartile range, PPO: preferred provider organization, EPO: exclusive provider organization, HMO: health maintenance organization, POS: point-of-service, CDHP: consumer directed health plan, HDHP: high deductible health plan

### Healthcare utilization

The number of insurance claims per year for SD patients (median: 0.51, IQR: 0.33-1.00) was greater than in SHL (median: 0.40, IQR: 0.25-0.67, p <0.001) and in AMD (median: 0.50, IQR: 0.32-1.00, p <0.001). The most common healthcare service utilized for SD patients was fundus photography (n=2,614, 11.98%). Medical examination and evaluation (n=57,950, 15.32%) was the most common healthcare service in AMD. Comprehensive audiometry threshold evaluation and speech recognition (n=308,619, 27.42%) was the most common healthcare service in SHL. Table 2 details the top five services utilized in each group.

**Table 2.** Most common healthcare services utilized

| Healthcare service/CPT code | N (%) |
| --- | --- |
| **SD** |  |
| Fundus photography (92250) | 2,614 (11.98) |
| Medical examination and evaluation (92014) | 2,343 (10.74) |
| Scanning computerized ophthalmic diagnostic imaging of the retina (92134) | 1,994 (9.14) |
| Fluorescein angiography (92235) | 1,329 (6.09) |
| Extended ophthalmoscopy (new visit or diagnosis) (92226) | 1,215 (5.57) |
| **SHL** |  |
| Comprehensive audiometry threshold evaluation and speech recognition (92557) | 308,619 (27.42) |
| Tympanometry (impedance testing) (92567) | 181,190 (16.10) |
| Tympanometry and reflex threshold measurements (92550) | 66,965 (5.95) |
| Office visit-established patient (99213) | 54,304 (4.83) |
| Office visit-new patient (99203) | 34,838 (3.10) |
| **AMD** |  |
| Medical examination and evaluation (92014) | 57,950 (15.32) |
| Fundus photography (92250) | 51,245 (13.55) |
| Scanning computerized ophthalmic diagnostic imaging of the retina (92134) | 50,508 (13.35) |
| Extended ophthalmoscopy (new visit or diagnosis) (92226) | 27,246 (7.20) |
| Fluorescein angiography (92235) | 21,472 (5.68) |

CPT: current procedural terminology, SD: stargardt disease, SHL: bilateral sensorineural hearing loss, AMD: non-exudative age related macular degeneration

### Healthcare costs

Payments per year for SD (median [IQR], $105.58 [$50.53-$218.71]) were greater than for SHL ($51.01 [$25.66-$121.66], p <0.001) and AMD ($76.20 [$38.00-$164.86], p <0.001). Table 3 presents the regression analysis for payments for each condition, adjusting for age, sex, type of benefit plan and year of first service.

**Table 3.** Multivariable quantile regression for payments per covered year

| **Stargardt disease** |  |  |
| --- | --- | --- |
| **Variables** | **Coefficient (95% CI)** | **P-value** |
| Age | -0.75 (-0.98, -0.53) | **0.001** |
| Sex |  |  |
| Male | Reference |  |
| Female | 8.82 (0.98, 16.67) | **0.03** |
| Year service was rendered |  |  |
| 2010 | Reference |  |
| 2011 | -29.61 (-40.90, -18.32) | **<0.001** |
| 2012 | -41.52 (-53.22, -29.81) | **<0.001** |
| 2013 | -26.83 (-38.91, -14.74) | **<0.001** |
| 2014 | -27.24 (-39.52, -14.96) | **<0.001** |
| Type of benefit plan |  |  |
| PPO | Reference |  |
| Comprehensive | 7.25 (-19.81, 34.31) | 0.599 |
| EPO | -6.22 (-27.79, 15.34) | 0.572 |
| HMO | -15.73 (-28.06, -3.40) | **0.012** |
| POS | -1.60 (-16.76, 13.56) | 0.836 |
| POS with capitation | 28.66 (-21.34, 78.66) | 0.261 |
| CDHP | 10.41 (-8.44, 29.26) | 0.279 |
| HDHP | 6.81 (-12.53, 26.15) | 0.490 |
| **Bilateral sensorineural hearing loss** |  |  |
| **Variables** | **Coefficient (95% CI)** | **P-value** |
| Age | -0.32 (-0.34, -0.31) | **<0.001** |
| Sex |  |  |
| Male | Reference |  |
| Female | -3.62 (-4.11, -3.12) | **<0.001** |
| Year service was rendered |  |  |
| 2010 | Reference |  |
| 2011 | -16.30 (-17.07, -15.53) | **<0.001** |
| 2012 | -20.54 (-21.31, -19.77) | **<0.001** |
| 2013 | -18.11 (-18.89, -17.32) | **<0.001** |
| 2014 | -12.65 (-13.43, -11.87) | **<0.001** |
| Type of benefit plan |  |  |
| PPO | Reference |  |
| Comprehensive | -1.87 (-3.41, -0.32) | **0.018** |
| EPO | 6.99 (5.45, 8.52) | **<0.001** |
| HMO | -4.17 (-4.93, -3.42) | **<0.001** |
| POS | -1.92 (-2.86, -0.98) | **<0.001** |
| POS with capitation | -1.35 (-4.49, 1.80) | 0.402 |
| CDHP | -2.61 (-3.75, -1.47) | **<0.001** |
| HDHP | 0.26 (-1.06, 1.58) | 0.701 |
| **Nonexudative age-related macular degeneration** |  |  |
| **Variables** | **Coefficient (95% CI)** | **P-value** |
| Age | 1.60 (1.44, 1.76) | **<0.001** |
| Sex |  |  |
| Male | Reference |  |
| Female | 3.03 (1.69, 4.36) | **<0.001** |
| Year service was rendered |  |  |
| 2010 | Reference |  |
| 2011 | -38.54 (-40.48, -36.59) | **<0.001** |
| 2012 | -48.88 (-50.87, -46.89) | **<0.001** |
| 2013 | -47.87 (-49.89, -45.84) | **<0.001** |
| 2014 | -51.77 (-53.82, -49.72) | **<0.001** |
| Type of benefit plan |  |  |
| PPO | Reference |  |
| Comprehensive | -14.68 (-18.04, -11.31) | **<0.001** |
| EPO | 1.97 (-2.23, 6.17) | 0.357 |
| HMO | -4.56 (-6.88, -2.23) | **<0.001** |
| POS | -0.62 (-3.05, 1.81) | 0.615 |
| POS with capitation | 10.02 (2.07, 17.96) | **0.014** |
| CDHP | -9.22 (-12.41, -6.03) | **<0.001** |
| HDHP | 1.72 (-2.11, 5.55) | 0.378 |

CI: confidence interval, PPO: preferred provider organization, EPO: exclusive provider organization, HMO: health maintenance organization, POS: point-of-service, CDHP: consumer directed health plan, HDHP: high deductible health plan

For SD patients, female gender was associated with higher annual payments as compared to males (p=0.03). However, older age (per year) was associated with lower payments of $0.75 per year. Annual payments varied by type of plan (HMO $15.73 lower than PPO, p=0.012).

For SHL, EPO plans were associated with higher payments per year ($6.99) than PPO plans (p<0.001). Factors associated with lower payments included higher age, female gender, year of first service in the database (2011-2014), and type of plan.

For AMD, older age (per year) was associated with an additional $1.60 payment (p<0.001). Payments for females were $3.22 higher than those for males (p<0.001). Those with POS with capitation plans incurred $10.02 more per year than those with PPO plans (p=0.014).

When comparing payments per year for SD to those for SHL, adjusted payments per year were significantly higher for SD patients (Table 4). The adjusted median payments for SD was $ 101.06 (95% confidence interval [CI]: $ 98.92-$ 103.20) and $53.23 (95%CI: $52.98-$53.48) for SHL.

**Table 4.** Payments per covered year for SD vs. SHL

| Variables | Coefficient (95% CI) | P-value |
| --- | --- | --- |
| Condition |  |  |
| SD | Reference |  |
| SHL | -47.83 (-49.98, -45.67) | <0.001 |
| Age | -0.33 (-0.34, -0.31) | <0.001 |
| Sex |  |  |
| Male | Reference |  |
| Female | -3.57 (-4.07, -3.07) | <0.001 |
| Year service was rendered |  |  |
| 2010 | Reference |  |
| 2011 | -16.40 (-17.17, -15.63) | <0.001 |
| 2012 | -20.67 (-21.44, -19.89) | <0.001 |
| 2013 | -18.21 (-18.99, -17.42) | <0.001 |
| 2014 | -12.76 (-13.55, -11.97) | <0.001 |
| Type of benefit plan |  |  |
| PPO | Reference |  |
| Comprehensive | -1.73 (-3.30, -0.17) | 0.030 |
| EPO | 6.81 (5.27, 8.35) | <0.001 |
| HMO | -4.23 (-4.99, -3.47) | <0.001 |
| POS | -1.93 (-2.88, -0.99) | <0.001 |
| POS with capitation | -1.13 (-4.30, 2.03) | 0.483 |
| CDHP | -2.54 (-3.69, -1.39) | <0.001 |
| HDHP | 0.28 (-1.05, 1.61) | 0.676 |

CI: confidence interval, SD: stargardt disease, SHL: bilateral sensorineural hearing loss, PPO: preferred provider organization, EPO: exclusive provider organization, HMO: health maintenance organization, POS: point-of-service, CDHP: consumer directed health plan, HDHP: high deductible health plan

The difference in payments per year was $47.83 (95% CI: $45.67 - $49.98, p<0.001). EPO plans were associated with higher payments per covered year.

Because of the age difference in SD and AMD patients, we also compared the AMD data with that of SD patients aged ≥50 years (n=1,669) (Table 5). A similar trend in the result was observed. The adjusted median payment for SD patients aged ≥50 years was $96.43 (95%CI: $91.35-$101.51) and was $79.09 (95%CI: $78.43-$79.74) for AMD. Therefore, payments for SD patients were $17.34 higher than for AMD (95% CI: $12.22 - $22.47, p<0.001). Factors associated with significantly greater payments included older age, female gender, and type of benefit plan (POS with capitation).

**Table 5.**
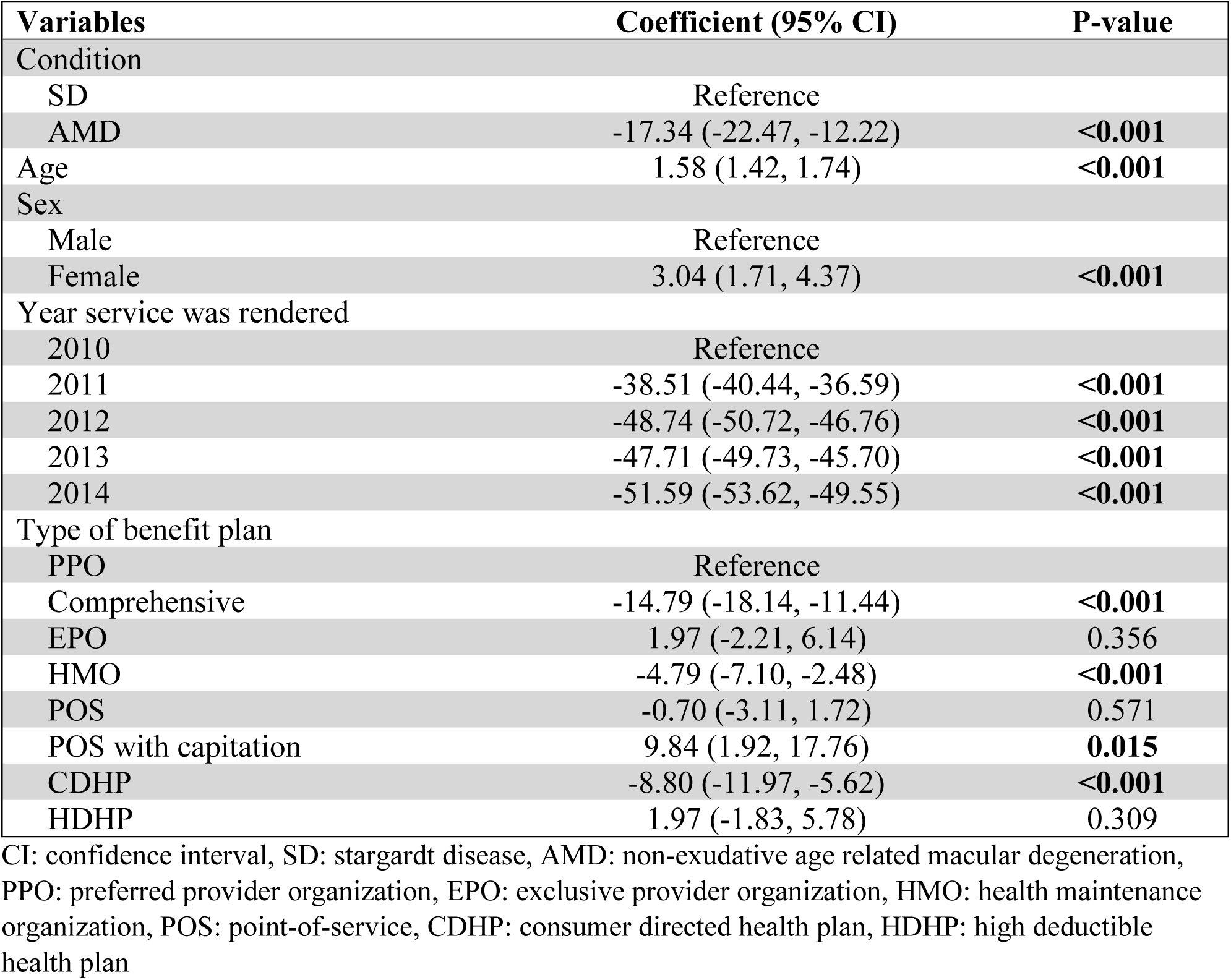
Payments per covered year for SD vs. AMD

## Discussion

The data show that there is a significant annual healthcare cost associated with SD. Payments associated with SD were higher than two control conditions, SHL and AMD. Payments also appeared to be influenced by patient gender, possibly due to differences in gender-related disease progression^12^ or health-seeking behavior^13^. To our knowledge, this is the first published estimate of the direct healthcare costs associated with SD.

The annual unadjusted median cost for patients with SD in our study, using only the MarketScan commercial claims and encounters database, was $105.58. A prior study estimated the healthcare costs of retinitis pigmentosa (RP), another inherited degenerative retinal disease, to be $3206 per patient^14^. This is higher than the figure reported by our study. This difference could be related to the fact that the RP study included both the MarketScan^®^ commercial claims and encounters and the Medicare supplemental datasets. Also, the RP study included claims with an ICD-9 diagnosis of retinitis pigmentosa in both the primary or secondary diagnosis position. In this study, we limited the analyzed claims to those with a primary diagnosis of SD. While limiting the dataset, this enhances the specificity of results to SD-related healthcare encounters. Future studies on the costs of SD could integrate multiple datasets to see if the SD-related costs are comparable to those of RP.

Greater awareness of the economic burden of SHL and AMD exists possibly because of their higher prevalence than SD. In one study which used Medicare data, the annual per-patient cost for dry AMD was reported to be $204.43^8^, a higher amount than we found. This difference could be due to the use of a different database, with an older population. A study from United Kingdom, including children aged 7 to 9 years with congenital bilateral hearing impairment, reported an annual per-patient healthcare cost of $2808, again higher than in our data. The study only included children with over 60 dB hearing loss, relied mostly on self-reported cost information from parents, included costs related to inpatient care and the use of community and social care services, as well as the cost of cochlear implants, hearing aids, and other assistive devices such as loop systems and special alarm clocks^15,16^.

One of the potential factors driving the high total healthcare costs for SD is the relatively early onset of disease. SD is usually diagnosed in the second or third decades of life and the associated visual loss is progressive thereafter. The total direct costs of diseases that present at an early age could be higher than diseases that present later, due to accumulation of costs over time. While this study does not address the impact of the disease on productivity and quality of life, one can extrapolate that a disease with onset at earlier ages would lead to a greater lifetime impact on productivity and quality of life than diseases that occur in late life. Vision impairment has a significant impact on education, employment, productivity, and independence of daily living. Vision impairment is also associated with falls, injuries, reduced mobility, depression, poor health outcomes, and reduced quality of life^17^. One study reported that the total economic burden, including direct and indirect costs, was $27.5 billion per year (2012 dollars) among people aged 40 years and younger with eye disorders^18^. Estimates of the total economic burden of SD, and its indirect costs, are not yet available.

This study has several limitations. First, the data were limited to patients younger than 65 years. Therefore, the overall costs might be under-reported for AMD. Second, the dataset used for the current study does not include Medicare and Medicaid data, and therefore these costs are not included. Third, individuals who are uninsured or unemployed were not captured in the MarketScan database, because it includes only employment-based claims. Fourth, since there is no treatment available for SD, a vast majority of the patients rely on low vision devices, especially in the later stages of the disease^19^. These low vision devices are not covered by many insurance companies in the United States and therefore the associated cost was not fully captured herein. Further, the nature of any administrative claims database is such that there is a possibility of diagnosis and procedure codes not being recorded in a standardized way. Lastly, disease stage or severity was not considered in this study.

Further research could explore other datasets, and include the costs attributable to low vision aids, guide dogs, loss of productivity, reduction in hours worked, and caregiver costs. It would also be useful to evaluate visual function and disease severity in relation to economic impact. The development of novel therapeutic modalities such as retinal stem cell and gene therapy^3,20–27^ are likely to affect the long-term economic burden associated with SD.

## Conclusion

In this limited dataset, the annual per-patient costs of SD were found to be higher than those incurred by patients with bilateral SHL and non-exudative AMD. The total lifetime per-patient cost of SD may exceed that of non-exudative AMD, due to the earlier age of onset of the former condition. There is a need to develop cost-effective treatments for SD, to reduce the long-term economic burden associated with this condition.

## Data Availability

The MarketScan data used in this project was purchased by the Johns Hopkins University and is covered by a Data Use Agreement that prohibits release of the data to the general public.

